# Predictive value of sudden olfactory loss in the diagnosis of COVID-19

**DOI:** 10.1101/2020.04.27.20081356

**Authors:** Antje Haehner, Julia Draf, Sarah Dräger, Katja de With, Thomas Hummel

## Abstract

**Introduction:** Recent reports suggest that sudden smell loss might be a symptom of SARS-CoV-2 infection. The aim of this study was to investigate the frequency of olfactory loss in an out-patient population who presented to a coronavirus testing center during a 2-week period and to evaluate the diagnostic value of the symptom “sudden smell loss” for screening procedures.

**Methods:** In this cross-sectional controlled cohort study, 500 patients who presented with symptoms of a common cold to a corona testing center and fulfilled corona testing criteria, completed a standardized diagnostic questionnaire which included the patients’ main symptoms, time course and an additional self-assessment of the patients’ current smell, taste function and nasal breathing compared to the level before onset of symptoms.

**Results:** Out of the 500 patients, 69 presented with olfactory loss. Twenty-two of them subsequently tested positive for SARS-CoV-2. Only twelve out of the patients without olfactory loss tested positive, resulting in a frequency of 64.7% for the symptom “sudden smell loss” in COVID-19 patients. Compared to COVID-19 patients without smell loss, they were significantly younger and less severely affected. Changes in nasal airflow were significantly more pronounced in SARS-CoV-2 negative patients with olfactory complaints compared to the patients with smell loss who were tested positive for SARS-CoV-2. By excluding patients with a blocked nose, the symptom “sudden smell loss” can be attested a high specificity (97%) and a sensitivity of 65% with a PPV of 63% and NPV of 97% for COVID-19.

**Conclusion:** Considering the high frequency of smell loss in non-hospitalized COVID-19 patients, acute olfactory impairment should be included in the WHO symptoms list and should be recognized as an early symptom of the disease. In contrast to other acute viral smell impairment, COVID-19 associated smell loss seems to be only rarely accompanied by a severely blocked nose.

## Introduction

Viral upper respiratory tract infections (URTI) are one of the most common causes of olfactory loss. Together with rhinoviruses, adeno-, influenza-, and parainfluenza viruses, coronaviruses have long been known to account for at least 70% of common colds [1]. Compared to other respiratory viruses however, the frequency of reported olfactory disorders in coronavirus infections was lower so far and has never been a serious health issue [2,3,4].

During an acute URTI, nasal congestion and rhinorrhea are commonly accompanied by a temporary smell loss of varying severity. Significant olfactory loss without rhinitic symptoms however, is comparatively rare and points towards damage of the olfactory epithelium. URTI-associated smell loss typically occurs after the fifth decade of life and becomes apparent to the patients several days after onset of the cold [5]. According to a number of anecdotal reports and very few published studies, a different pattern is seen in patients with severe acute respiratory syndrome coronavirus 2 (SARS-CoV-2) infection. Smell and/or taste loss have been observed in 5-80% of these patients [6,7], sometimes as the only apparent symptom, at a younger age, and a very early stage of the disease. The diagnostic accuracy of this symptom is controversially discussed considering the fact that more than 200 viruses are known to cause URTIs with a subsequent olfactory loss.

The aim of this study was to investigate the frequency of olfactory loss in an out-patient population who presented to a coronavirus testing center during a 2-week period and to evaluate the diagnostic value of this symptom for screening procedures.

## Material and Methods

All patients who presented to the coronavirus testing center at the University hospital Dresden routinely received a standardized diagnostic questionnaire which included the patients’ main symptoms, time course and an additional self-assessment of the patients’ current smell, taste function and nasal breathing compared to the level before onset of symptoms. The patients had to indicate whether they experienced loss of smell and/or taste (yes vs. no). For quantifying olfactory/gustatory function and nasal breathing, a visual analogue scale (VAS) with its extreme left of the scale defined as “no function” (0 units), and the extreme right of the scale defined as highest function possible (“extremely good”- 10 units) was used.

Out of 620 patients who fully completed the diagnostic questionnaire, 500 met the criteria for SARS-CoV-2 testing and throat swabs were collected according to WHO recommendations [8]. In 34 patients (6.8%), the diagnosis of SARS-CoV-2 infection was confirmed by means of a positive RT-PCR test.

## Results

### Study population

Out of the 500 patients (mean age, 41.3 years; range, 18–86 years) who presented with symptoms of a common cold (mean duration, 5.8 days; range, 1–32 days) and fulfilled corona testing criteria, 45.4% were male and 54.6% female. Main symptoms were: cough (76.8%), sore throat (64.8%), rhinorrhea (55.2%), myalgia (40.0%), dyspnea (33.4%), and fever (20.8%). Sixty-nine patients (13.8%) reported sudden smell and/or taste loss.

### COVID-19 patients

Out of the 34 patients who were tested positive for SARS-CoV-2, 22 (64.7%) complained about sudden smell and/or taste loss. The chemosensory loss started 1–2 days before other symptoms in one patient, at the same time in four patients and 1–7 days after in 14 patients; 3 patients were not sure about the smell loss onset. COVID 19 patients with olfactory loss were significantly younger than COVID-19 patients without smell loss (p=0.04) and had less severe symptoms (Table 1). Smell loss in COVID 19 patients was characterized by a severe loss of function from an average of *8.8* to *2* and minor changes in nasal breathing from an average of *8.4* to *7.3*, as rated on the 10 point VAS scales. Only 14 patients complained about rhinorrhea.

**Table 1.** Patient characteristics. COVID-19 patients with (correct) and without olfactory loss (false negative) and SARS-CoV-2-negative-patients with smell loss (false positive) out of a total patient population of n=500

|  | **correct**<br>**(n=22)** | **false negative**<br>**(n=12)** | **false positive**<br>**(n=47)** |
| --- | --- | --- | --- |
| Gender female | 54.5% | 33.3% | 65.2% |
| Age (M±SD), years | 38.6 ± 12.4 | 47.9 ± 10.8 | 39.3±12.4 |
| Symptoms onset before testing (M±SD), days | 5.5 ± 3.2 | 4.3 ± 2.9 | 7.0 ± 4.9 |
| Smell/taste loss onset before testing (M±SD), days | 3.1 ± 3.3 |  | 5.4 ± 3.8 |
| Smell VAS score difference (M±SD) | -6.8 ± 2.3 | 0 | -5.3 ± 1.9 |
| Nasal breathing VAS score difference (M±SD) | -1.1 ± 1.6 | -0.6 ± 0.9 | -4.9 ± 2.4 |
| Cough | 81.0% | 75.0% | 91.5% |
| Sore throat | 42.9% | 58.3% | 83.0% |
| Dyspnea | 19.0% | 25.0% | 59.6% |
| Fever | 19.0% | 58.3% | 23.4% |
| Rhinorrhea | 45.5% | 33.3% | 80.9% |
| Myalgia | 38.1% | 50.0% | 55.3% |

While only one patient complained about isolated taste loss, the majority of patients reported either combined smell and taste loss, or smell loss only.

### Patients with smell loss without SARS-CoV-2 infection

Four hundred sixty six patients were tested negative for SARS-CoV-2, 47 out of them (10.1%) reported disorder-associated smell loss. Olfactory impairment started 1–14 days after the other symptoms had begun and was characterized by both severe loss of function (mean value, *8.8* to *3.4*) and impaired nasal breathing (mean value, *8.5* to *3.6*) as rated on the 10-point VAS scale. The majority of these patients (80.9%) suffered from rhinorrhea (Table 1).

Disease-associated changes in nasal airflow were significantly more pronounced in the 47 SARS-CoV-2 negative patients with olfactory complaints compared to the 22 patients with smell loss who were tested positive for SARS-CoV-2 (p<0.001) (Figure 1). In contrast, changes in olfactory function were rated more severe in the latter group (p=0.023).

**Figure 1.**
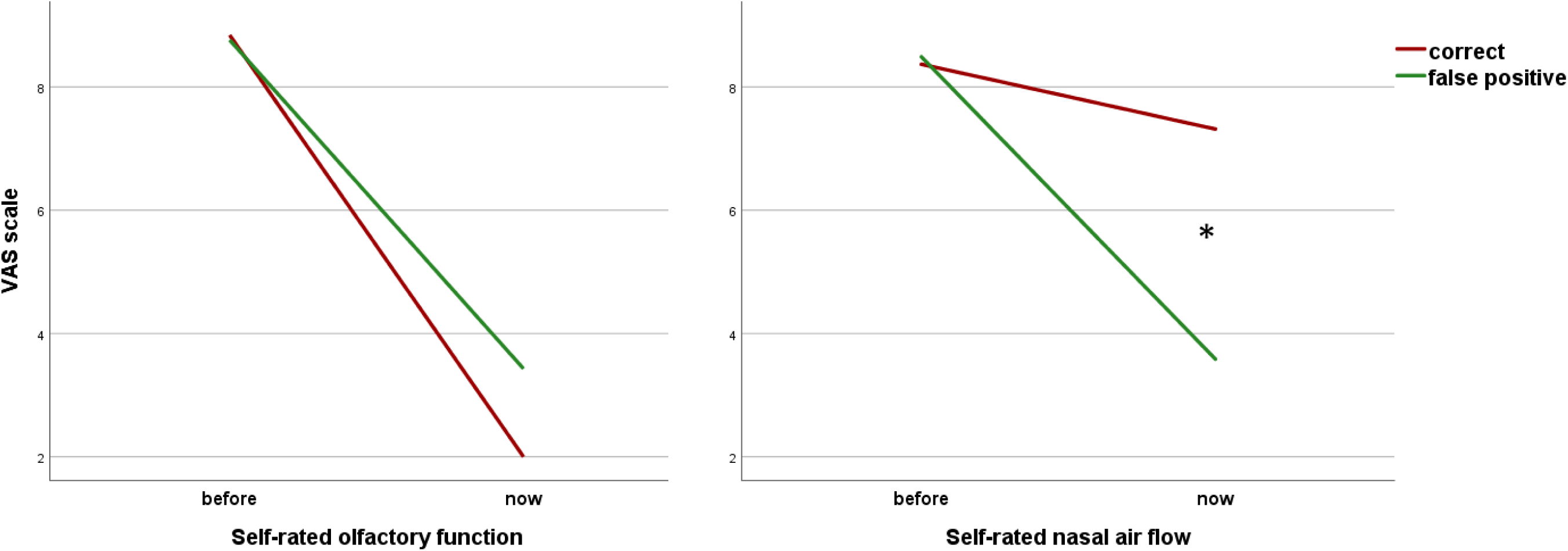
Change in self-rated olfactory function and nasal breathing compared to the level before disease in SARS-CoV-2-positive (correct) and in SARS-CoV-2-negative-patients with smell loss (false positive)

### Predictive value (PV)

Using our data of an out-patient population, the sole symptom “smell loss” had the following diagnostic characteristics: 65% sensitivity, 90% specificity, 32% positive predictive value (PPV), and 97% negative predictive value (NPV). Taking into account the low frequency of sinunasal symptoms in COVID-19 patients and excluding those with a blocked nose from the false positive sample however, positive PV increased to 63% and specificity to 97%.

## Discussion

In this large URTI patient cohort study of a coronavirus testing center, we found that 13% of patients reported sudden olfactory loss. Out of these patients, 32% were subsequently tested positive for SARS-CoV-2. On the other hand, 2.8% of the patients without olfactory loss were tested positive.

In this out-patient sample, 64.7% of COVID-19 patients observed smell and/or taste loss early or even before other symptoms started. Compared to COVID-19 patients without smell loss they were significantly younger and less severely affected. In contrast to other acute viral smell impairment, COVID-19 associated smell loss seems to be only rarely accompanied by a severely blocked nose and thus the effect on olfaction may be independent of nasal congestion. By excluding patients with a blocked nose, the symptom “sudden smell loss” can be attested a high specificity (97%) and sensitivity of 65% with a PPV of 63% and NPV of 97% for COVID-19.

Thus, considering the high frequency of smell loss in non-hospitalized COVID-19 patients, acute olfactory impairment should be included in the WHO symptoms list and should be recognized as an early symptom of the disease. It also suggests the need for an exhaustive olfactory and gustatory follow-up of these patients in order to assess the extent of the olfactory loss and the significance of the chemosensory loss in terms of the prognosis of the patients.

## Data Availability

Data are available upon request.

## Statement of Ethics

Investigations were performed according to the Guidelines for Biomedical Studies Involving Human Subjects (“Helsinki Declaration”). The protocol was approved by the Ethics Committee of the Medical Faculty of the TU Dresden (EK-115032020) and all subjects provided written informed consent.

## References

1. Heikkinen T, Jarvinen A. The common cold. Lancet 2003;361:51–9.

2. Sugiura M, Aiba T, Mori J, Nakai Y. An epidemiological study of postviral olfactory disorder. Acta Otolaryngol Suppl. 1998;538:191–6.

3. Konstantinidis I, Haehner A, Frasnelli J, Reden J, Quante G, Damm M, et al. Post-infectious olfactory dysfunction exhibits a seasonal pattern. Rhinology 2006;44(2): 135–9.

4. Suzuki M, Saito K, Min WP, Vladau C, Toida K, Itoh H, et al. Identification of viruses in patients with postviral olfactory dysfunction. Laryngoscope 2007;117(2):272–7.

5. Hummel T, Whitcroft KL, Andrews P, Altundag A, Cinghi C, Costanzo RM, et al. Position Paper on Olfactory Dysfunction. Rhinol Suppl. 2017;54(26):1–30.

6. Mao L, Wang M, Chen S, Hu Y, Chen S, He Q, et al. Neurological manifestations of hospitalized patients with COVID-19 in Wuhan, China. JAMA Neurol. 2020 Apr 10. doi: 10.1001/jamaneurol.2020.1127. [Epub ahead of print]

7. Lechien JR, Chiesa-Estomba CM, De Siati DR, Horoi M, Le Bon SD, Rodriguez A, et al. Olfactory and gustatory dysfunctions as a clinical presentation of mild-to-moderate forms of the coronavirus disease (COVID-19): a multicenter European study. Eur Arch Otorhinolaryngol. 2020 Apr 6. doi: 10.1007/s00405-020-05965-1. [Epub ahead of print]

8. https://www.who.int/emergencies/diseases/novel-coronavirus-2019/technical-guidance. Assessed April 6, 2020

